# Higher testosterone and testosterone/estradiol ratio in men are associated with better epigenetic estimators of mortality risk

**DOI:** 10.1101/2023.02.16.23285997

**Authors:** Cynthia DJ Kusters, Kimberly C Paul, Ake T Lu, Luigi Ferrucci, Beate R Ritz, Alexandra M Binder, Steve Horvath

**Author notes:** **Corresponding author:** Cynthia DJ Kusters. Department of Epidemiology, Fielding School of Public Health at UCLA, 650 Charles E. Young Drive South, Box 708822 Los Angeles, CA 90095-7088, USA.

## Abstract

**Introduction:** Sex hormones are hypothesized to drive sex-specific health disparities. Here, we study the association between sex steroid hormones and DNA methylation-based (DNAm) biomarkers of age and mortality risk including Pheno Age Acceleration (AA), Grim AA, and DNAm-based estimators of Plasminogen Activator Inhibitor 1 (PAI1), and leptin concentrations.

**Methods:** We pooled data from three population-based cohorts, the Framingham Heart Study Offspring Cohort (FHS), the Baltimore Longitudinal Study of Aging (BLSA), and the InCHIANTI Study, including 1,062 postmenopausal women without hormone therapy and 1,612 men of European descent. Sex hormone concentrations were standardized with mean 0 and standard deviation of 1, for each study and sex separately. Sex-stratified analyses using a linear mixed regression were performed, with a Benjamini-Hochberg (BH) adjustment for multiple testing. Sensitivity analysis was performed excluding the previously used training-set for the development of Pheno and Grim age.

**Results:** Sex Hormone Binding Globulin (SHBG) is associated with a decrease in DNAm PAI1 among men (per 1 standard deviation (SD): -478 pg/mL; 95%CI: -614 to -343; P:1e-11; BH-P: 1e-10), and women (-434 pg/mL; 95%CI: -589 to -279; P:1e-7; BH-P:2e-6). The testosterone/estradiol (TE) ratio was associated with a decrease in Pheno AA (-0.41 years; 95%CI: -0.70 to -0.12; P:0.01; BH-P: 0.04), and DNAm PAI1 (-351 pg/mL; 95%CI: -486 to -217; P:4e-7; BH-P:3e-6) among men. In men, 1 SD increase in total testosterone was associated with a decrease in DNAm PAI1 (-481 pg/mL; 95%CI: -613 to -349; P:2e-12; BH-P:6e-11).

**Conclusion:** SHBG was associated with lower DNAm PAI1 among men and women. Higher testosterone and testosterone/estradiol ratio were associated with lower DNAm PAI and a younger epigenetic age in men. A decrease in DNAm PAI1 is associated with lower mortality and morbidity risk indicating a potential protective effect of testosterone on lifespan and conceivably cardiovascular health via DNAm PAI1.

**Graphic abstract:** Visual representation of our main results stratified by sex
There were four outcomes of interest in the rectangular shapes in the middle of this figure, Pheno-Age Acceleration (AA), Grim AA, DNAm-based PAI1, and DNAm-based leptin. We measured five hormone concentrations (testosterone, estrone, estradiol, DHEAS, and Sex Hormone Binding Globulin (SHBG)). In addition, one hormone level ratio (testosterone / estradiol) was estimated. Associations were calculated by linear mixed regression models between sex hormones and the outcomes of interests. The associations are represented by colored arrows with the lines’ thickness representing the strength of the association. As the association was measured mainly cross-sectional, the directionality of the association cannot be established. Hormone levels were inversely associated with epigenetic estimators of mortality risk.
Abbreviations: E1: total estrone; E2: total estradiol; SHBG: Sex Hormone Binding Globulin; TotT: total testosterone; TE ratio: Total testosterone divided by total estradiol concentration

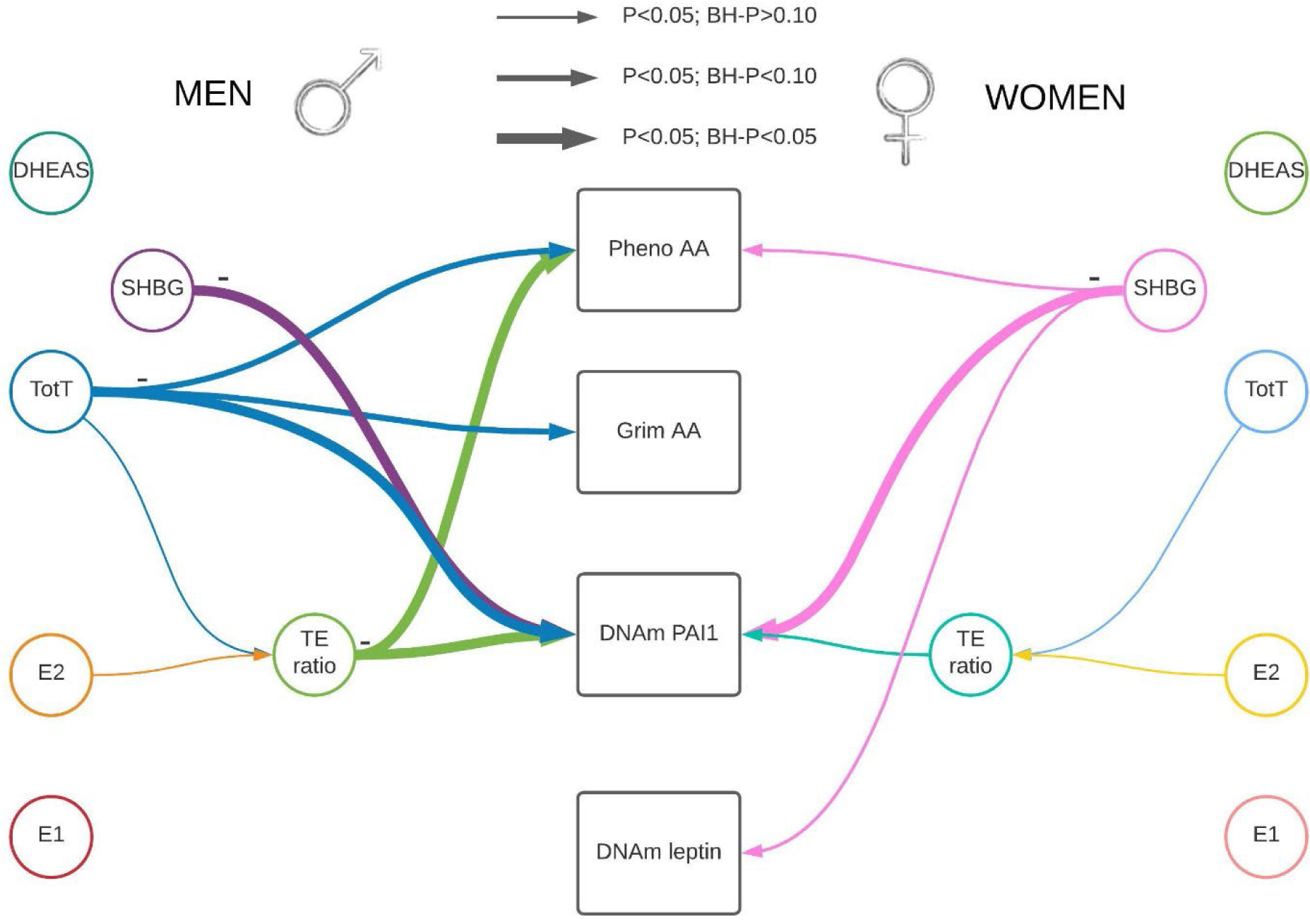

## Introduction

Sex hormones are hypothesized to drive sex-specific health disparities.^1^ Females are overrepresented among the elderly population, and experience lower mortality rates and cardiovascular disease (CVD) risk.^2, 3^ Prior studies suggest women may also have a higher risk of some neurodegenerative diseases.^3^ A higher concentration of various sex hormones concentrations has been hypothesized to decrease mortality and morbidity.^1, 4^

Steroid sex hormones are a group of hormones that play a role in sexual development and reproduction, although they have many additional functions. Three main categories are estrogens, progesterone, and androgens. Estradiol and estrone are estrogens and female-dominated sex hormones while androsterone sulfate, androstenedione, dehydroepiandrosterone sulfate (DHEAS), and testosterone are androgens and considered male-dominated sex hormones. A testosterone/estradiol (TE) ratio can be used as a proxy for hormonal balance between androgens and estrogens. This ratio has also been associated with various health outcomes, including cerebrovascular disease, metabolic disorders and CVD.^5–7^ Sex hormone binding globulin (SHBG) is a protein that transports sex hormones, binds estrone, estradiol and testosterone, and influences bioavailability of these hormones.^8^

The sex steroid hormones have time-trends. Among adult women, major changes of sex hormones concentrations occur during menopause. Around menopause, estradiol (and estrone to a more limited degree) decreases rapidly over 6 months, then slowly decreases over time with the largest decrease in the first 3 years. After menopause, estrone become the major estrogen.^9^ Among postmenopausal women, testosterone and androsterone also decrease slowly with age, while SHBG appears to change with age in a U-shaped form, with lowest concentrations around 60-70 years of age, followed by an increase with age.^9–12^ For men, SHBG increases over time,^11–13^ and estradiol remains stable or decreases slightly,^14, 15^ and testosterone decrease over time.^12, 13, 15^ DHEAS peaks around 20-24 years of age among men and women and then decrease with age.^13, 16–18^

Animal studies indicate that higher estrogen concentrations are associated with a longer lifespan among male mice, but not in females.^19–21^ Observational human studies suggest that low androgen levels among men, and possibly postmenopausal women, are associated with CVD and their risk factors.^22, 23^ Among women, a majority of the studies have used female reproductive characteristics as a proxy for hormone levels, as sex hormones increase after menarche and estradiol, a strong estrogen, decreases after menopause. Several studies have indicated that women who gave birth later in life (>40 years), had high parity, or late menopause with longer exposure to higher levels of estrogens had lower all-cause mortality rates,^24–31^ while in contrast high levels of postmenopausal estradiol concentrations have been associated with an increased mortality.^32^

Biomarkers of aging that are strongly associated with increased morbidity and mortality can serve as surrogate endpoints for these outcomes. Specifically, these indicators of biological age can be used to investigate whether an exposure of interest might be associated with morbidity and mortality risk, though causality cannot be established. Recently epigenetic age has become a widely used indicator of biological age, as it has been shown to be strongly associated with morbidity and mortality.^33–35^ Interestingly, among centenarians, epigenetic (and hence biologic) age has been shown to be lower.^36^ There are some indications that sex hormones influence epigenetic age and that lower sex hormone concentrations (due to earlier age of menopause, ovariectomy, or lower ovarian reserve etc.) could accelerate epigenetic aging.^37–40^ Among the various epigenetic age measures, GrimAge accelerations (AA) and Pheno AA are strongly associated with mortality and morbidity, especially CVD.^35^ GrimAge is established by a two-step approach that combines DNA methylation (DNAm) based estimates of plasma protein levels, DNAm based estimates of smoking pack years, age and sex into a mortality risk estimate. Of these DNAm based estimates of protein biomarkers, plasminogen activator inhibitor 1 (PAI1) is most strongly associated with morbidity and leptin shows sex differential effects.^35^

PAI1 is a protein that is involved in tissue hemostasis and an increase in PAI1 is associated with metabolic syndrome, lipid metabolism, and cardiovascular health.^41–45^ Genetic mutations in the SERPINE1 mutation, associated with PAI-1 concentrations, have been associated with cellular senescence, leukocyte telomere length, and longevity.^46^ Previously, studies that assessed associations between sex hormones and PAI1 protein concentrations in blood reported conflicting results. Among men, several small studies indicated that higher testosterone and SHBG concentrations are associated with lower PAI1 concentrations.^47, 48^ SHBG is also inversely correlated with PAI1 concentrations among women.^49^ Conversely, there is some indication that higher testosterone and DHEAS in women may be associated with higher PAI1 concentrations, though results from two studies were not consistent.^49, 50^

Leptin is a peptide hormone and is associated with regulation of food intake and energy balance. Leptin also influences inflammatory processes, angiogenesis, lipolysis, and neuroplasticity.^51–53^ For women, several studies indicated positive correlations between estrone, estradiol and testosterone and leptin concentrations,^54–58^ whereas reported correlations of leptin with SHBG and DHEAS were inconsistent.^55–58^ Body mass index (BMI) is strongly positively associated with leptin concentration as leptin is secreted by adipocytes.^15^ In addition, adipose tissue can convert estrone to estradiol.^15, 59^ Hence, BMI could be a strong confounder. Adjusting for BMI attenuated the correlations between sex hormones (estradiol, testosterone, SHBG and DHEAS) and leptin concentrations in some studies.^56, 57^ Among men, higher testosterone concentrations are correlated with lower leptin concentrations.^57, 58, 60–62^ Two studies among men indicated a negative association between SHBG and leptin concentration,^58, 61^ one no association,^60^ and yet another study a positive association.^57^ These inconsistencies might be due to sample size, differences in population characteristics (age, ethnicity etc.), or due to methodological differences.

In this study, we are focusing on the association between five sex steroid hormones (estrone, estradiol, SHBG, testosterone, DHEAS), the testosterone/estradiol (TE) ratio, and epigenetic age acceleration for older men and postmenopausal women. We chose to restrict the study to postmenopausal women without hormone therapy as female sex hormones stabilize after menopause,^9^ and assessment of sex hormones is not influenced by timing within the menstrual cycle. We combined data from three large studies that have collected DNAm as well as measured sex hormones in the blood of men and women. No previous studies have reviewed whether sex hormones are also associated with the epigenetic biomarkers for leptin (DNAm Leptin) and PAI1 (DNAm PAI1). In addition, no previous studies on sex hormone concentrations and epigenetic based age acceleration have been performed. We hypothesize that higher concentrations of sex hormones are associated with lower epigenetic age.

### Main results

Our study included 1,612 men and 1,062 postmenopausal women of European descent, who were not using hormone therapy at the time of blood draw, with at least one sex hormone measurement and genome-wide DNA methylation (DNAm) array data. Estrone was only available in the Framingham Heart Study (FHS), while DHEAS was available in the Baltimore Longitudinal Study of Aging (BLSA) and the InCHIANTI.

The average age at sex hormones assay was 66.5 years (standard deviation [SD]: 9.8) for women and 62.4 years for men (SD: 12.2). The time between the sex hormone measurement and subsequent blood collection for DNAm analysis among women and men of the FHS study was 6.6 and 6.6 years (SD: 0.6, SD: 0.7, for women and men respectively). For subjects from the InCHIANTI and BLSA, sex hormones and DNAm were measured during the same visit. Sex hormones for women were measured on average 17.4 years (SD: 10.7) after their menopause with an average age of menopause of 49.0 years (SD: 5.9). See more details on the characteristics in Table 1, and the characteristics stratified by study including the sex hormone concentrations in supplemental Table 1. The average concentrations for total sex hormones were comparable with previous studies of individuals of similar age.^63–65^ Due to differences in sex hormone concentrations and differences in characteristics between the three studies, as well as different methods used to estimate sex hormones concentration, the sex hormones were standardized (mean 0; standard deviation 1) for each sex within their study, followed by winsorization of the top and bottom 1% (Supplemental Table 1). See supplemental figure 1 for scatterplots of the sex hormone concentration by age and study.

**Table 1.** Overview of the characteristics of the study population.

|  |  | **Women** |  |  | **Men** |  |  |
| --- | --- | --- | --- | --- | --- | --- | --- |
|  |  | **Mean / N** | **SD / %** | **N** | **Mean / N** | **SD / %** | **N** |
| **Characteristics** |  |  |  |  |  |  |  |
| Study | FHS | 665 | 62.6 | 1062 | 1,088 | 67.5 | 1612 |
|  | InCHIAN TI | 193 | 18.2 |  | 222 | 13.8 |  |
|  | BLSA | 204 | 19.2 |  | 302 | 18.7 |  |
| Age at sex hormone measurement | years | 66.5 | 9.8 | 1062 | 62.4 | 12.2 | 1612 |
| Age at DNA methylation | years | 70.6 | 8.9 | 1062 | 66.9 | 11.6 | 1612 |
| Time between visits among FHS | years | 6.6 | 0.6 | 665 | 6.6 | 0.7 | 1088 |
| Age of menopause | years | 48.6 | 6.0 | 991 | NA |  |  |
| Time since menopause at sex hormone measurements | years | 17.3 | 11.0 | 991 | NA |  |  |
| Smoking status at sex hormone measurements | never | 437 | 40.7 | 1075 | 485 | 29.9 | 1624 |
|  | Former/current | 638 | 59.3 |  | 1,139 | 70.1 |  |
| Packyears at sex hormone measurements among smokers | pack-years | 17.8 | 18.4 | 638 | 24.1 | 20.8 | 1139 |
| Alcohol intake | Drinks/weeks | 7.0 | 9.2 | 1015 | 19.4 | 24.2 | 1488 |
| BMI at sex hormone measurement | kg/m2 | 27.5 | 5.2 | 1073 | 28.3 | 4.4 | 1622 |
| **Hormone levels** |  |  |  |  |  |  |  |
| Total estrone | pg/ml | 32.2 | 20.9 | 634 | 50.6 | 17.7 | 1037 |
| Total estradiol | pg/ml | 11.0 | 19.6 | 984 | 23.9 | 10.1 | 1519 |
| SHBG | nmol/L | 83.3 | 49.4 | 832 | 61.7 | 31.5 | 1260 |
| Total testosterone | ng/dl | 35.4 | 28.0 | 1017 | 541.3 | 226.6 | 1533 |
| DHEAS | ug/dl | 86.2 | 71.4 | 192 | 143.1 | 115.6 | 221 |
| **Bioavailable sex hormones** |  |  |  |  |  |  |  |
| Bioavailable estrone | pg/ml | 1.0 | 0.7 | 633 | 1.8 | 0.6 | 1037 |
| Bioavailable estradiol | pg/ml | 0.2 | 0.3 | 812 | 0.5 | 0.2 | 1250 |
| Bioavailable testosterone | pg/ml | 4.0 | 3.0 | 829 | 82.7 | 35.0 | 1259 |
| **Main outcome** |  |  |  |  |  |  |  |
| Pheno AA | years | -0.7 | 6.4 | 1075 | 0.8 | 6.1 | 1624 |
| Grim AA | years | -1.6 | 4.0 | 1075 | 1.7 | 4.5 | 1624 |
| DNAm PAI1 | pg/ml | 19021 | 2862 | 1075 | 20865 | 3117 | 1624 |
| DNAm Leptin | pg/ml | 12026 | 2839 | 1075 | 4769 | 1648 | 1624 |

Abbreviations: N: Number; SD: Standard Deviation; years: years; NA: Not applicable; FHS: Framingham Heart Study; BLSA: Baltimore Longitudinal Study of Aging

### Correlations

Pairwise correlations were calculated and represented in supplemental figure 2. The correlations between the five sex hormones (estrone, estradiol, SHBG, total testosterone, and DHEAS) varied between 0.01 and 0.66. While the correlation between total estrone and SHBG was relatively weak (r=0.10 in men; r=0.01 in women), the correlation between estrone and estradiol was relatively high (r=0.59 in men; r=0.66 in women). The correlations between Pheno AA, Grim AA, DNAm PAI1, and DNAm leptin, i.e., the outcomes we will focus on as the strongest surrogate markers for morbidity and mortality, were weak to moderate in size, i.e., between 0.06 and 0.45 (supplemental figure 3).

### Sex hormones and main outcomes (Grim AA, Pheno AA, DNAm PAI1 and DNAm leptin)

Each sex hormone concentrations were standardized within sex and study group. The standardized sex hormone concentrations were pooled over the three studies for males and females separately. We used linear mixed regression models to analyze the association between sex hormones and the epigenetic clocks/DNAm proteins to take family relatedness within the FHS into account. We also adjusted for age, BMI, average alcohol intake, smoking packyears, physical activity, time between visits, cohort study, and blood cell composition based on DNAm. For all postmenopausal women, we also included time since menopause at the time sex hormones were measured. All analyses were performed sex stratified and we adjusted for multiple testing (Number of tests in the Benjamini Hochberg analysis: 24). Our main findings for the association between sex hormones and epigenetic clocks/DNAm proteins are summarized in table 2, figures 1 & 2, with the precursors for total and bioavailable sex hormones displayed in Figure 2.

**Table 2.** Linear mixed regression analysis with random intercept were performed to assess the effect of sex hormones on Pheno Age, GrimAge, DNAm-based PAI1, and DNAm-based leptin. Sex hormones were standardized and winsorized (highest and lowest 1%) by study. The analyses were performed stratified by sex, and adjusted for age, body mass index (BMI), alcohol intake, smoking packyears, physical activity, the time between visits, cohort study, and blood cell composition based on DNAm. Among women, we also adjusted for time since menopause.

| Outcome | Sex Hormones | Men |  |  |  |  | Females |  |  |  |  |
| --- | --- | --- | --- | --- | --- | --- | --- | --- | --- | --- | --- |
|  |  | Est. | 95% LL | 95% UL | P | BH-P | Est. | 95% LL | 95% UL | P | BH-P |
| Pheno AA | E1 | 0.26 | -0.09 | 0.61 | 0.14 | 0.31 | 0.06 | -0.42 | 0.54 | 0.79 | 0.96 |
|  | E2 | 0.13 | -0.15 | 0.40 | 0.37 | 0.67 | 0.05 | -0.32 | 0.41 | 0.81 | 0.96 |
|  | SHBG | -0.11 | -0.41 | 0.19 | 0.47 | 0.67 | -0.40 | -0.75 | -0.04 | 0.03 | 0.22 |
|  | TotT | -0.35 | -0.64 | -0.06 | 0.02 | 0.08 | 0.11 | -0.23 | 0.44 | 0.54 | 0.92 |
|  | TE ratio | -0.41 | -0.70 | -0.12 | 0.01 | 0.04 | -0.12 | -0.48 | 0.23 | 0.49 | 0.92 |
|  | DHEAS | -0.02 | -0.62 | 0.57 | 0.94 | 0.98 | 0.38 | -0.12 | 0.87 | 0.14 | 0.47 |
| Grim AA | E1 | 0.19 | -0.02 | 0.39 | 0.07 | 0.22 | 0.03 | -0.22 | 0.29 | 0.80 | 0.96 |
|  | E2 | -0.04 | -0.21 | 0.12 | 0.59 | 0.71 | 0.04 | -0.15 | 0.23 | 0.66 | 0.96 |
|  | SHBG | 0.05 | -0.13 | 0.22 | 0.59 | 0.71 | -0.10 | -0.29 | 0.08 | 0.28 | 0.76 |
|  | TotT | -0.22 | -0.39 | -0.05 | 0.01 | 0.06 | 0.00 | -0.18 | 0.18 | 0.99 | 0.99 |
|  | TE ratio | -0.16 | -0.33 | 0.01 | 0.07 | 0.22 | -0.14 | -0.32 | 0.05 | 0.16 | 0.48 |
|  | DHEAS | 0.28 | -0.08 | 0.64 | 0.12 | 0.30 | 0.09 | -0.16 | 0.34 | 0.50 | 0.92 |
| DNAm PAI1 | E1 | 61 | -100 | 222 | 0.46 | 0.67 | -67 | -270 | 136 | 0.52 | 0.92 |
|  | E2 | -103 | -230 | 24 | 0.11 | 0.30 | 139 | -22 | 299 | 0.09 | 0.37 |
|  | SHBG | -478 | -614 | -343 | 1E-11 | 1E-10 | -434 | -589 | -279 | 1E-07 | 2E-06 |
|  | TotT | -481 | -613 | -349 | 2E-12 | 6E-11 | -134 | -284 | 15 | 0.08 | 0.37 |
|  | TE ratio | -351 | -486 | -217 | 4E-07 | 3E-06 | -197 | -356 | -39 | 0.02 | 0.18 |
|  | DHEAS | 68 | -200 | 335 | 0.62 | 0.71 | -21 | -264 | 222 | 0.86 | 0.96 |
| DNAm LEPTIN | E1 | 0 | -90 | 89 | 1.00 | 1.00 | 13 | -162 | 187 | 0.89 | 0.96 |
|  | E2 | -28 | -101 | 45 | 0.44 | 0.67 | 28 | -106 | 163 | 0.68 | 0.96 |
|  | SHBG | -30 | -109 | 50 | 0.46 | 0.67 | -140 | -271 | -10 | 0.04 | 0.22 |
|  | TotT | -26 | -103 | 52 | 0.52 | 0.69 | 42 | -82 | 166 | 0.50 | 0.92 |
|  | TE ratio | 18 | -60 | 95 | 0.66 | 0.72 | -26 | -159 | 107 | 0.70 | 0.96 |
|  | DHEAS | -75 | -253 | 104 | 0.41 | 0.67 | 9 | -174 | 193 | 0.92 | 0.96 |

Abbreviations: Est: Effect estimate; LL: Lower Limit; UL: Upper Limit; P: P-value; BH-P: Benjamini-Hochberg adjusted P-value; AA: Age Acceleration; DNAm: DNA-methylation; E1: total estrone; E2: total estradiol; SHBG: Sex Hormone Binding Globulin; TotT: total testosterone; TE ratio: Total testosterone divided by total estradiol concentration

**Figure 1.**
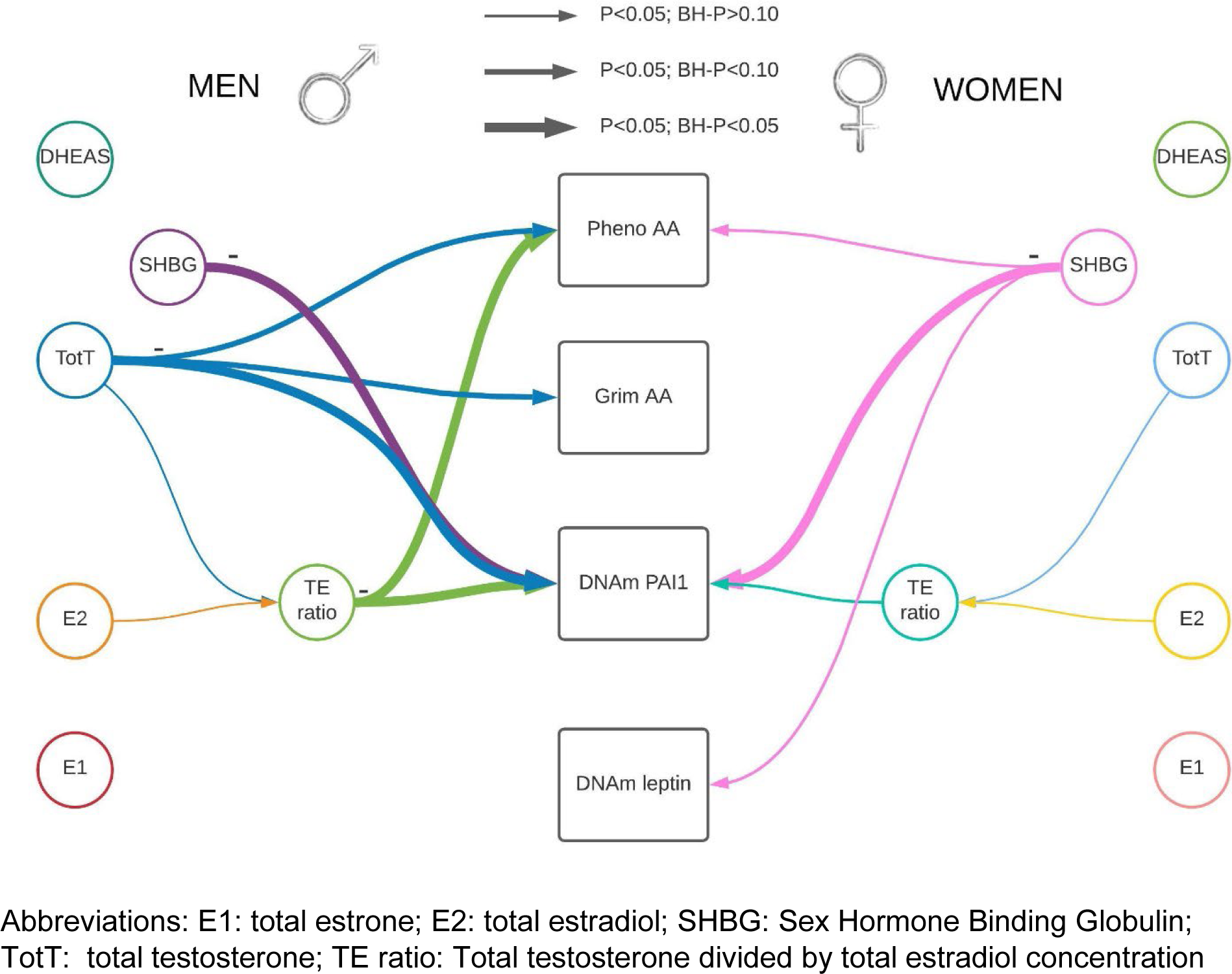
Visual representation of our main results stratified by sex. There were four outcomes of interest in the rectangular shapes in the middle of this figure, Pheno-Age Acceleration (AA), Grim AA, and DNAm-based PAI1 and DNAm-based leptin. We measured five hormone concentrations (testosterone, estrone, estradiol, DHEAS, and Sex Hormone Binding Globulin (SHBG)). In addition, one hormone level ratio (testosterone / estradiol) was estimated. Associations identified in a linear mixed regression model are represented by colored arrows between sex hormones and the outcomes of interests, with the lines’ thickness representing the strength of the association. Hormone levels were inversely associated with epigenetic estimators of mortality risk.

**Figure 2.**
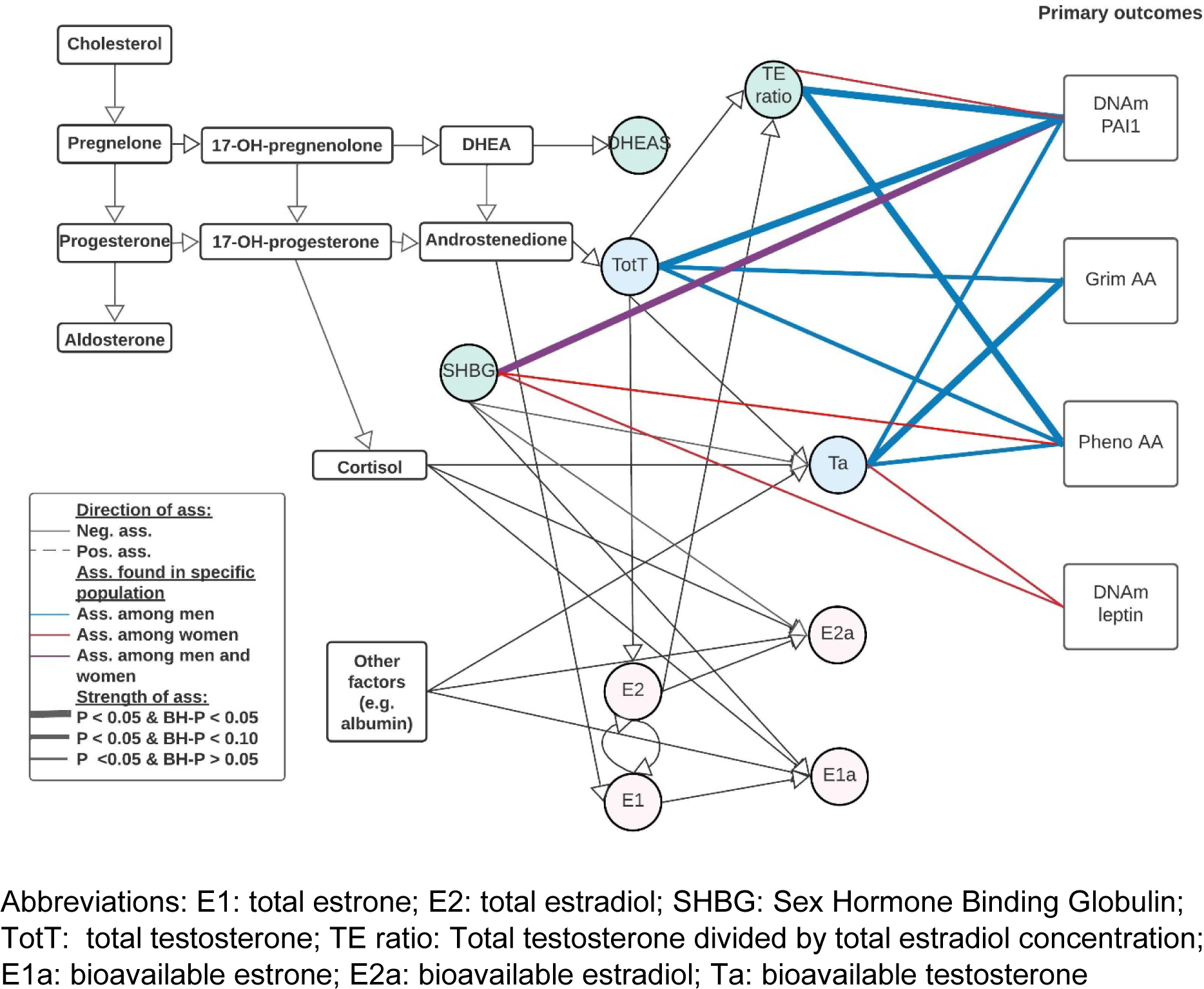
Visual representation of our main results including bioavailable testosterone and precursors. There were 4 outcomes of interest in the rectangle shapes on the right, Grim Age Acceleration (AA), Pheno AA, and DNAm-based PAI1, and DNAm-based leptin. There were 8 various hormone levels, 5 total concentrations of testosterone, estrone, estradiol, DHEAS, and SHBG, and 3 bioavailable concentrations of testosterone, estrone and estradiol. In addition, there was one hormone level ratio (testosterone / estradiol) estimated. Associations were estimated using a linear mixed regression model, please note that these are not causal estimates but associations only.

In line with our hypothesis that high sex hormone concentrations are associated with better epigenetic age or epigenetic profile, among males, we identified four significant associations all indicating a negative association. None of these associations significantly differed by cohort (Supplemental table 2). Three of these associations were with DNAm PAI1, namely SHBG, total testosterone, and the TE ratio. The fourth was an association between the TE ratio and Pheno AA. A one SD increase in SHBG was associated with lower DNAm PAI1 by -478 pg/mL, which is equivalent to approximately 0.17 SD of DNAm PAI1 (95%CI: -614 to -343; P-value: 1e-11; Benjamini Hochberg adjusted P-value (BH-P): 1e-10). One SD increase in total testosterone was associated with a similar decrease in DNAm PAI1 (-481 pg/mL; 95%CI: -6213 to -349; P-value: 2e-12; BH-P: 6e-11), and one SD increase in the TE ratio decreased DNAm PAI1 by 351 pg/ml (95%CI: -620 to -355; P-value: 2e-12; BH-P: 4e-11). An increase of 1 SD in TE ratio was associated with a decrease in Pheno AA with 0.41 years (95%CI: -0.70 to -0.12; P-value: 0.01; BH-P: 0.04); i.e., suggesting a “younger” DNAm Pheno age compared to chronological age. For two additional hormones there was a decrease in epigenetic age acceleration suggested (P-value < 0.05 or BH-P < 0.10); specifically, an increase in total testosterone appeared to decrease both Grim AA and Pheno AA. With each increase of 1 SD in testosterone Grim AA decreases by 0.22 years (95%CI: -0.39 to -0.05; P-value: 0.01; BH-P: 0.06), and Pheno AA by 0.35 years (95%CI: -0.64 to -0.06 years; P-value: 0.02; BH-P: 0.08). When adjusting for DNAm PAI1 in the model for the effect of testosterone on Grim AA, the association between testosterone and Grim AA attenuated (0.03 yrs; 95%CI: -0.13 to 0.19 years; P-value: 0.71), whereas adjusting for Grim AA in the model between testosterone and DNAm PAI1, the estimated effect remained the same (-413 pg/ml; 95%CI: -534 to -291; P-value: 6e-11).

Among females, only SHBG was statistically significantly associated with DNAm PAI1, and three suggestive associations were detected, that is even though the P-value was less than 0.05, the BH-P was above 0.10. One SD increase of SHBG was associated with a decrease of DNAm PAI1 (-434; 95%CI: 589 to -279; P-value: 1e-7; BH-P: 2e-6). There was indication for heterogeneity based on the likelihood ratio test adding the interaction term for SHBG and study (P:4e-3). In addition, an increase in the TE ratio was associated with a decrease in the DNAm PAI1 (-197 (95%CI: -356 to -39; P-value: 0.02; BH-P: 0.18), and an increase in SHBG was associated with a decrease in Pheno AA by 0.40 years (95%CI: -0.75 to -0.04; P-value 0.03; BH-P: 0.22). Finally, SHBG suggestively decreased DNAm leptin by approximately 140 pg/ml per SD increase of SHBG (95%CI: -271 to -10; P-value: 0.04; BH-P: 0.22).

### Combined outcomes

We combined our four main outcomes (Grim AA, Pheno AA, DNAm PAI1 and DNAm leptin) into one summarized outcome by normalizing (mean:0, and SD: 1) each outcome and averaging the overall outcome (see table 3). This analysis indicated that an increase in SHBG, testosterone, and TE-ratio were associated with a decreased summarized outcome (SHBG: -0.05 SD; 95%CI: -0.08 to -0.02; P-value: 2e-3; BH-P: 4e-3; testosterone: -0.07 SD; 95%CI: -0.10 to -0.04; P-value: 4e-6; BH-P: 3e-5; and TE-ratio: -0.05 SD; 95%CI: -0.08 to -0.02; P-value: 1e-3; BH-P: 4e-3). Among females, an increase in SHBG was associated with a decrease in our overall outcome (-0.08 SD; 95%CI: -0.11 to -0.04; P-value: 9e-5; BH-P: 5e-4). We also created a latent variable using PCA, and the results were almost identical (see supplemental table 3).

**Table 3.**
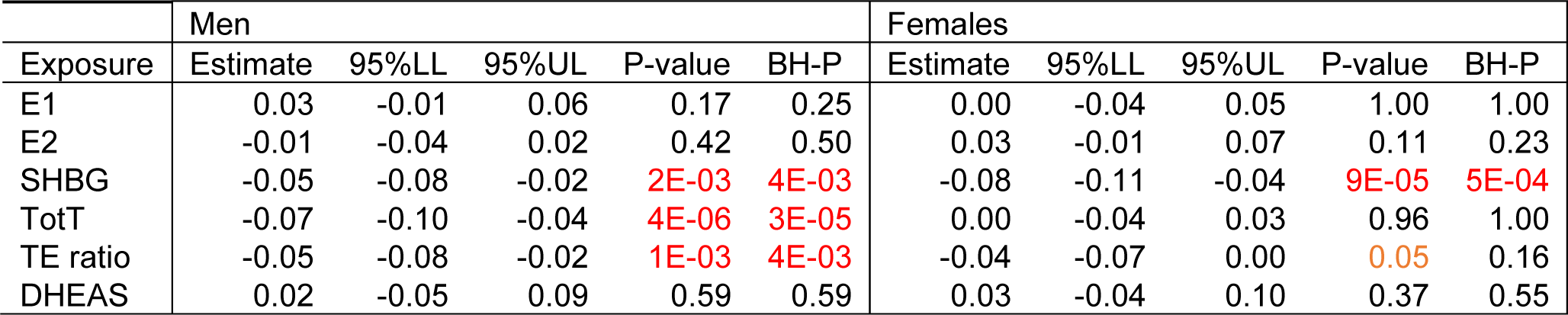
Association between sex hormones and an aggregate outcome. We normalized each outcome first (mean zero; standard deviation one) followed by summarization of the four outcomes (Pheno AA, Grim AA, DNAm PAI1, and DNAm Leptin) into one aggregate. Associations were calculated using a linear mixed model adjusting for familial relationships.

|  | Men |  |  |  |  | Females |  |  |  |  |
| --- | --- | --- | --- | --- | --- | --- | --- | --- | --- | --- |
| Exposure | Estimate | 95%LL | 95%UL | P-value | BH-P | Estimate | 95%LL | 95%UL | P-value | BH-P |
| E1 | 0.03 | -0.01 | 0.06 | 0.17 | 0.25 | 0.00 | -0.04 | 0.05 | 1.00 | 1.00 |
| E2 | -0.01 | -0.04 | 0.02 | 0.42 | 0.50 | 0.03 | -0.01 | 0.07 | 0.11 | 0.23 |
| SHBG | -0.05 | -0.08 | -0.02 | 2E-03 | 4E-03 | -0.08 | -0.11 | -0.04 | 9E-05 | 5E-04 |
| TotT | -0.07 | -0.10 | -0.04 | 4E-06 | 3E-05 | 0.00 | -0.04 | 0.03 | 0.96 | 1.00 |
| TE ratio | -0.05 | -0.08 | -0.02 | 1E-03 | 4E-03 | -0.04 | -0.07 | 0.00 | 0.05 | 0.16 |
| DHEAS | 0.02 | -0.05 | 0.09 | 0.59 | 0.59 | 0.03 | -0.04 | 0.10 | 0.37 | 0.55 |

### Bioavailable sex hormones

While our main analysis focused on the total concentration of estrone, estradiol and testosterone, we also calculated the bioavailable concentrations of these sex hormones relying on the mass action law calculation.^66^ Bioavailable sex hormones are the concentration of the sex hormone that are considered bioactive and have the possibility to be utilized in biologic processes. The bioavailable and total concentrations of sex hormones were strongly correlated, and were strongly correlated with SHBG, which is one of the main binding proteins of sex hormones. The correlations between the total and bioavailable sex hormone concentrations ranged between 0.68 and 0.98 (see supplemental figure 2, supplemental table 4).

Results for all sex hormones, including bioavailable sex hormones, are available in supplemental table 5. Most of the results for bioavailable sex hormones were very similar to those that were seen for the total concentration of the specific sex hormone. This was expected due to the high correlation between total and bioavailable sex hormone concentration.

However, some differences were observed, bioavailable estradiol was suggestively associated with an increase in DNAm PAI1 in men and women (men: 110; 95%CI: -21 to 240; P-value: 0.10; BH-P: 0.28 / women: 220; 95%CI: 57 to 382; P-value: 0.01; BH-P: 0.15). Among men, a slightly stronger association, compared to total testosterone, between bioavailable testosterone and Grim AA was identified (1 SD increase in bioavailable testosterone was associated with a decrease of 0.28 years in Grim AA (95%CI: -0.46 to -0.11; P-value: 2e-3; BH-P: 0.01)). On the contrary, the effect for DNAm PAI1 was attenuated for bioavailable versus total testosterone (-160; 95%CI: -299 to -22; P-value: 0.02; BH-P: 0.09) among men, and appeared to be attenuated or even change directions among women (-134; 95%CI: -284 to 15; P-value: 0.08, BH-P: 0.31; and 142; 95%CI: -10 to 294; P-value: 0.07, BH-P: 0.31, for total and bioavailable testosterone, respectively). Finally, there appears to be a positive association between bioavailable testosterone and DNAm leptin among females, though this was not statistically significant after correction for multiple testing (134; 95%CI: 8 to 260; P-value: 0.04, BH-P: 0.23).

### Other epigenetic AA and sex hormones

We additionally studied the acceleration measures from 6 epigenetic clocks (IEAA, EEAA, Horvath AA, Hannum AA, Skin-blood clock AA, and Dunedin PoAM; see supplemental table 6).^67–70^ There were no statistically significant associations between epigenetic age acceleration and sex hormones after adjusting for multiple testing. The strongest associations were seen among men for the TE ratio, with a decrease in IEAA (-0.35; 95%CI: -0.60 to -0.11; P-value: 5e-3, BH-P: 0.11) and Horvath AA (-0.37; 95%CI: -0.61 to -0.12; P-value: 3e-3, BH-P: 0.11).

### Sensitivity analysis

Using non-winsorized data instead of winsorized data, most of the effect estimates remained very similar (see supplemental table 7). InCHIANTIThe cross-sectional (BLSA & InCHIANTI) and prospective (FHS) effect estimates were comparable among males (supplemental table 8). Among females, there appeared to be some differences. Cross-sectionally, estradiol was associated with an increase in DNAm PAI1, while no association was seen in the longitudinal, prospective FHS study. SHBG and testosterone were associated with a more strongly negative association (decrease) in DNAm PAI1 in the FHS study, while the analysis based on the cross-sectional studies indicated a smaller effect size estimate for SHBG, and possibly a null or positive association for total testosterone.

A subset of FHS was used for the creation of the original GrimAge clock, as well as the DNAm biomarkers used for the GrimAge clock,^35^ while InCHIANTI was used for the creation of Pheno age.^33^ We expect the DNAm-based estimates to have a much stronger correlation with their estimand in the training set, allowing us to capture more of the effect of the estimand (e.g., plasma PAI1) on sex hormone levels. There was however no indication for heterogeneity by study. When excluding the training sets (so excluding InCHIANTI for Pheno AA and excluding the individuals from the training set in the FHS for Grim AA, DNAm PAI1, and DNAm leptin), the sample size reduced considerably to 869 females and 1390 males for the Pheno AA; and 569 females and 793 males for the other outcomes. Among this subset, our primary findings among males remained significant, with some associations slightly attenuated (supplemental table 9).

We also gathered a group of 90 male and 86 female individuals of Black race/ethnicity (BLSA) for our sensitivity analysis. We performed the same analysis for this subgroup (see supplemental table 10) and we found similar effect estimates. Nevertheless, studies with larger sample sizes are needed to replicate our findings and ensure that they are indeed valid for Black and other race/ethnicities.

## Discussion

We found that an increase in testosterone and the TE ratio in men was associated with a decreased epigenetic age acceleration, suggesting that higher testosterone levels are associated with biologically younger age and a lower DNAm-based estimate of PAI1 concentration. In both men and women, an increase in SHBG was associated with a lower DNAm-based PAI1 concentration. These findings may indicate that SHBG in both sexes and testosterone in men influence the morbidity, especially cardiovascular risks, and mortality risk in members of these population-based cohorts (BLSA, FHS, and InCHIANTI).

Increases in PAI1 concentration have been positively associated with risk for CVD, ischemic heart disease and stroke, with metabolic abnormalities such as diabetes and metabolic syndrome, and increased inflammation.^45, 71^ While the correlation between plasma PAI-1 and DNAm PAI-1 was only moderate (r=0.36), DNAm PAI-1 was shown to be a better surrogate for lifespan than the actual plasma measure, and performs better than Grim AA regarding associations with the comorbidity-index.^35^ Another potential benefit of using DNAm-based biomarkers instead of plasma biomarkers is that the DNAm-based biomarkers are representing a longer average estimate of the biomarker concentration and are not as affected by day-to-day variations that could bias the results.^72–74^ In the study by Lu et al, a higher concentration of DNAm PAI-1 was strongly associated with coronary heart disease, hypertension, type 2 diabetes, computed tomography based measurements of adiposity, and early age of menopause for women, while lower DNAm PAI-1 was associated with disease free status and better physical functioning.^35^ In our study, we identified a negative association of DNAm PAI1 with total testosterone, TE ratio (men only), and SHBG. The association between testosterone with plasma PAI1 concentration has been previously described for measured (not DNAm based) PAI1 protein concentrations.^47, 48^ A recent review has described the current state of research findings on testosterone concentrations and CVD and vascular aging.^75^ Though PAI1 was not specifically reviewed, previously studies appeared to provide contradictory results for testosterone and CVD among women, while among men low testosterone concentration was associated with an increase in CVD and vascular aging.^75^ This is in line with our findings, where testosterone is associated with lower DNAm-based concentrations of PAI1, suggestive of a more protective cardiovascular profile. As the TE ratio is composed of testosterone and estradiol, it is not surprising that results are highly correlated with the total testosterone concentration. However, the balance represented by the TE ratio has been shown to have an influence on many health outcomes, and were found to be associated with libido, cognitive function, well-being and mental state, increased muscle mass, loss of fat mass, increased basal metabolic rate, and reduced cardiovascular markers.^76^

With regards to the association between SHBG and DNAm PAI1 concentration, an increase in SHBG concentration was associated with an on average decrease in bioavailable hormone concentrations (estrone, estradiol, testosterone). Total testosterone and SHBG were both associated with a decrease in DNAm PAI1 concentration. This could indicate a direct effect of SHBG acting as a hormone, or be due to the fact that both SHBG and PAI1 are synthesized within hepatocytes.^8, 77–80^ One of the major factors that influence SHBG levels is high body mass index, or obesity, and even though we adjusted for this in our analysis, we cannot rule out residual confounding. In addition, other factors that influence SHBG, such as chronic infections, thyroid disease, and certain medications^81, 82^ could partially explain this finding. However, our findings do indicate that an increase in SHBG, and among men also total testosterone, is independently associated with a decrease in DNAm PAI1. Hence, SHBG, as well as testosterone among men may decrease PAI1 levels, and thereby positively influence the risk of developing CVD, metabolic disorders, and inflammation.

Among men only, testosterone and the TE ratio are both associated with a decrease in epigenetic age acceleration. Slightly stronger associations were found for Pheno AA, while the association of testosterone with Grim AA disappeared after taking DNAm PAI1 into account, suggesting that the effect of testosterone on Grim AA was driven by changes in the DNAm PAI1 concentrations. Previous studies that investigated the associations of testosterone with mortality or CVD reported contradictory and inconclusive results. Some early studies of testosterone replacement treatment and/or higher testosterone concentrations among men and possibly women suggested an increased risk for mortality or CVD.^83–89^ Other clinical trials and observational studies found a decreased mortality or morbidity when testosterone was increased.^90–94^ Two more recent, large studies reported testosterone to be associated with a decrease in overall mortality and cardiovascular-related deaths among men, but possibly an increase in women.^95, 96^ Thus, the increased mortality and CVD risk may be specific to men with low testosterone levels and these studies also suggested that the effect might not be linear. Yet, our own findings among men were not driven by those with low testosterone levels and we saw a linear effect for our measures. While our study did not review the association between testosterone and mortality directly, our results indicate that testosterone and the balance between testosterone and estradiol (TE ratio) is associated with an epigenetic profile predictive of lower mortality and lower risk for CVD among men. In addition, we also found some evidence for an association between the TE ratio and other epigenetic age acceleration clocks, e.g., for Horvath AA. Since the Horvath pan tissue clock was designed as an age estimator it is referred to as a first-generation clock. By contrast, GrimAge and PhenoAge are referred to as second generation clocks since they were designed as mortality risk predictors. Using this terminology, we find that the beneficial effect of the TE ratio in men can be detected using both first- and second-generation epigenetic clocks. As such, we consider it a highly robust finding. We did not find an association between sex hormones and epigenetic age acceleration among women. This could be due to multiple reasons. First, it could reflect that epigenetic clocks perform differently across the sexes. We think this explanation is unlikely since both GrimAge and PhenoAge predict mortality risk in both sexes. Second, it might be due to the relatively smaller sample size for women as we had to restrict our sample to postmenopausal women without hormone therapy to avoid confounding due to menopausal stage or hormone treatment. Third, it might be that sex hormone concentrations among postmenopausal women, except for SHBG, tend to generally be very low and their variability too limited to estimate effects reducing the power to identify associations in women.

Two of the three cohorts we used provided cross-sectional data, while among participants of the FHS, there was a significant time delay between the measurement of the sex hormones and the blood draw used for DNA methylation analyses. Most sex hormone concentrations decline with age, and it is possible that this resulted in an underestimation of effects in the FHS. However, most of the associations were consistent in the FHS and BLSA, especially for DNAm PAI1, while effect estimation in the InCHIANTI study alone appeared attenuated or null. Differences in estimated effect size across studies may also have been caused by differences in measurement techniques, relatively lower concentrations of sex hormones in the study, or by residual confounding from differences in lifestyle across the cohorts. The BLSA cohort is a multi-ethnic population among a highly educated population in an urban location. This population was relatively older than the other two at time of the assessment. The InCHIANTI study consists of relatively younger men (average age 61 years) at DNA methylation assessment and is a longitudinal study among subjects of Italian descent. As such, this population has a very different lifestyle then the other two studies. We adjusted for many potential confounders (including blood cell types, body mass index, smoking packyears, alcohol intake, physical activity, age at sex hormone assessment, time between measurements, study and for women time since menopause). Effect estimates varied slightly depending on the selection of confounders, with the strongest attenuation observed when we adjusted for BMI, which was expected to be a major confounder. However, given the differences in population characteristics, it is still possible that differences in lifestyle characteristics resulted in residual confounding.

The FHS was used in the original development of GrimAge (and hence also DNAm PAI1 and DNAm Leptin) and the InCHIANTI was used for the development of PhenoAge. As such, using individuals who were used in the training set for the development of the biomarkers could potentially introduce stronger correlation with the outcome (measured biomarkers and time to death). Our sensitivity analysis excluding these individuals indicated no significant difference in the effect estimates and association, suggesting that this potential bias did not change our overall results.

Our primary analysis consisted of men and women of Caucasian or European descent in order to avoid confounding by race/ethnicity. Thus, it is plausible that the results cannot be generalized to other races/ethnicities. However, our findings for a subgroup with Black ancestry subjects indicate that the association between total testosterone and epigenetic age accelerations as well as with DNAm PAI1 are similar at least among men (see also supplemental table 10). Further evaluation in larger minority populations or populations with different lifestyles, should be undertaken to confirm or refute our findings.

### Conclusion

A higher testosterone and a higher TE ratio, among men are associated with a decreased epigenetic age acceleration, suggesting a protective effect of testosterone. In addition, we found a strong effect of testosterone and TE ratio on the DNAm PAI1 concentration among men, and between SHBG and DNAm PAI1 concentration independent of sex, suggesting that both SHBG and testosterone, as well as the TE ratio (testosterone and TE ratio among men only) is associated with DNAm PAI1 and therefore potentially with cardiovascular health and overall mortality.

## Methods

### Study population

We used data from three population-based cohorts, the Framingham Heart Study (FHS), the Baltimore Longitudinal Study of Aging (BLSA) and the InCHIANTI Study. For more details regarding the study populations, see the supplemental table 1 and the supplemental note. Both FHS and InCHIANTI consist of subjects with European descent, while the BLSA has a more diverse population with 69.8% of European descent, 26.3% of African American or Black ancestry, and 3.9% of other descent. Most female individuals were postmenopausal (N: 1,504, 84.6%). In our main analysis, we restricted to postmenopausal women without hormone therapy and men of European descent.

### Sex hormones

For specific details about the measurement of sex hormones, we refer to the supplemental note. For sensitivity analysis, we calculated bioavailable estrone, estradiol and testosterone by using the mass action law when estimates of the total sex hormone (estrone, estradiol or testosterone) and SHBG were available.^66^ (Bioavailable) estrone was only available in the FHS, and DHEAS was only available in the InCHIANTI and BLSA.

Sex hormone concentrations were standardized with mean 0 and standard deviation of 1, for each study, sex, and postmenopausal status separately. We winsorized the outliers for sex hormone measurements by replacing the top and bottom outliers with the 1^st^ and 99^th^ percentile estimates, respectively. The winsorization was done stratified by sex, and study. Sex hormones were measured at the same time as blood DNA methylation for individuals in the BLSA and InCHIANTI study. For participants of the FHS, sex hormones were measured one visit prior to DNA methylation (average difference between visits: 6.6 years), the length of time between the measurements was added as a covariate in the model to take timing into account. For analysis, we removed postmenopausal women who reported that they were using hormone therapy at either the time of sex hormone or DNAm assessment (N=421), leaving 1,062 postmenopausal women without HRT.

### Epigenetic age accelerations and DNAm-based biomarkers

DNA methylation was performed using the Illumina 450k array on whole blood of all individuals in this study. For details regarding DNA methylation, we refer to the supplemental note. The epigenetic age accelerations and DNAm-based biomarkers were calculated using the DNAmage-website (https://dnamage.genetics.ucla.edu). We winsorized the top and bottom outliers (1%) for epigenetic age-accelerations and DNAm biomarkers, stratified by sex, and study. For this specific analysis, we focused on Pheno Age Acceleration (Pheno AA), Grim Age Acceleration (Grim AA), DNAm Plasminogen Activator Inhibitor-1 (PAI1) and DNAm leptin, as these have been shown to be the strongest epigenetic biomarker predictors for mortality and morbidity.^35^

### Statistical analysis

We performed our analysis, stratified by sex, among postmenopausal women without hormone therapy and all men. We analyzed the associations between various sex hormones and our outcomes using linear mixed regression analysis with a random intercept term to account for familial relationships from pedigrees in the FHS. We adjusted for age, body mass index (BMI), average alcohol intake, and smoking packyears estimated at the visit when sex hormones were assessed. We also adjusted for physical activity (in quartiles), the time between visits, cohort study, and blood cell composition based on DNAm. The estimated cell types (plasma blasts, exhausted T cells, naïve CD8 cells, CD4 T-cells, Natural Killer (NK) cells, monocytes, and granulocytes) were calculated using the Houseman and Horvath methods.^67, 97^ For all postmenopausal women, we also included time since menopause at the time sex hormones were measured. If confounders were missing (0.1% missing BMI, 5.3% missing physical activity, 7.3% missing alcohol intake), they were imputed using the MICE R-package. We adjusted for multiple testing by calculating the Benjamini-Hochberg adjusted P-value (BH-P) for both men and women separately (total of 24 tests per sex).

### Sensitivity analysis

Various sensitivity analyses were performed. These included analyzing bioavailable sex hormones with the outcomes, using non-winsorized data, stratifying by study, removing individuals from the original training set for the epigenetic outcomes, and analyzing the effect estimates among African American/Black participants only. Furthermore, we reviewed the influence of confounder selection. In addition to our main outcomes, we also performed linear mixed regression analysis for the association between the various sex hormones and six additional epigenetic age accelerations clocks.^67–70^

## Acknowledgments/Funding sources

This study was funded by NIH grants F32AG063442 (CK); K01AG072044 (KCP); U01AG060908 (CK; BR). This study was supported in part by the Intramural Research Program of the National Institute on Aging, NIH. The InCHIANTI study was supported as a “targeted project” (ICS 110.1/RS97.71) by the Italian Ministry of Health and by the U.S. National Institute on Aging (contracts N01-AG-916413, N01-AG-5-0002, and N01-AG-821336, and grant R01-AG-027012). AMB was supported by the National Cancer Institute of the National Institutes of Health under Award Number K07CA225856. The content is solely the responsibility of the authors and does not necessarily represent the official views of the National Institutes of Health.

## Supporting information

Supplemental figures descriptions

## Data Availability

All data produced in the present study are available upon request through each individual study

