## Supplemental note for "Higher testosterone and testosterone/estradiol ratio in men are associated with better epigenetic estimators of mortality risk"

**Supplemental note. Additional information about the study population**

Framingham Heart Study (FHS)

The Framingham Heart Study (FHS) is a prospective cohort study, initially focusing on cardiovascular disease. FHS began in 1948 with the recruitment of the Original cohort, recruiting 5,209 men and women between the ages of 30 and 62 from Framingham, Massachusetts.[1] In 1971, the children of the Original cohort (and the children’s spouses) were recruited within the study as a separate cohort (Offspring cohort). A sample of 5,124 men and women was recruited and followed prospectively.[2]

Participants from the FHS Offspring Cohort were eligible if they attended both the seventh and eighth examination cycles and had previously consented for the usage of molecular data. We used the 2,356 participants from the group of Health/Medical/Biomedical (IRB, MDS) consent and available for both Immunoassay array DNA methylation array data.

Peripheral blood samples, collected during the 8^th^ exam, was used for the analyses. DNA methylation quantification was conducted in two batches using the Illumina Infinium HumanMethylation450 array (Illumina). Methylation beta values were generated using the Bioconductor minfi package with Noob background correction.[3]

Using serum samples, the sex hormones estimates for testosterone and SHBG (males and females), and estrone and estradiol (males only) were performed using LC-MS/MS methods by the Boston Medical Center. The inter-assay cross validation (CV) for testosterone was between 3.5 and 7.8%; for estrone 9.6%, and estradiol 13.9%. Serum SHBG levels were measured using an immunofluorometric assay (DELFIA-Wallac, Inc., Turku, Finland). The inter-assay CVs were between 7.9% and 10.9%. Estrone and estradiol (females only) were measured by the Mayo clinic using the LC-MS/MS measured with inter-assay CV of 13.2% for estrone, 10.4% for estradiol.

For men, the lowest concentrations measured for these sex hormones were 4.03 pg/ml, 1.065 pg/ml, 7.6 nmol/L, and 12.7 ng/dl for estrone, estradiol, SHBG, and testosterone, respectively. For women, the lowest concentrations measured for these sex hormones were 5 pg/ml, 3 pg/ml, 9.1 nmol/L, and 1.7 ng/dl for estrone, estradiol, SHBG, and testosterone, respectively.

Among the FHS offspring cohort study, the population was mainly of European descent. Two individuals reported other race and were excluded from the analysis. Information about any of the sex hormones and DNA methylation was available for 1088 males and 1266 females. Among the females, 161 females were pre-menopausal at time of the sex hormone assessment. We excluded premenopausal women and those who reported use of hormone therapy at either visit 7 or visit 8 (N=416) or were missing information on hormone therapy at either visit (N=18). This left 665 postmenopausal women without hormone therapy for the analysis.

Baltimore Longitudinal Study of Aging (BLSA)

The Baltimore Longitudinal Study of Aging (BLSA) is American’s longest-running scientific study of aging. It originated in 1958.[4] This observational study measures physical and cognitive changes over time. BLSA participants are seen between 1- and 4-years intervals, depending upon their age at the visit.[5]

In BLSA, blood samples were collected for DNA extraction from 793 participants, and genome-wide DNA methylation was completed using the Illumina 450K. Background subtraction was applied using the preprocess Illumina command in the minfi Bioconductor package. Duplicate samples were randomly removed from the analysis.

Samples from the BLSA plasma bank were used to estimate the testosterone, SHBG, DHEAS and estradiol concentrations. Serum samples were drawn between 07:00 and 9:30am after fasting overnight. Duplicate estimates of testosterone and estradiol were estimated using aliquots of 100 µL serum, using ^125^I, double antibody radioimmunology (RIA) kits. The kits were obtained from Diagnostic Systems Laboratories, Inc (Webster, TX). SHBG concentrations were measured in 50 μL aliquots using RIA kits purchased from Radim (Liege, Belgium) which employ ^125^I labeled SHBG and PEG-complexed second antibody.[6,7] The analyses were done at the Covance Laboratories, Inc (Vienna, VA, USA).

For men, the lowest concentrations measured for these sex hormones were 1.5 pg/ml, 6.935 nmol/L, 5.4 ng/dl, and 16 ug/dl for estradiol, SHBG, testosterone, and DHEAS respectively. For women, the lowest concentrations measured for these sex hormones were 1.2 pg/ml, 14 nmol/L, 3 ng/dl, and 16 ug/dl for estradiol, SHBG, testosterone, and DHEAS, respectively.

Among the BLSA, the population was more diverse with 207 individuals reporting as Black race, and 30 individuals as other race. These individuals were excluded from the main analysis, although we did perform a sensitivity analysis on the individuals who reported as Black race. There were 793 individuals of European/White race. Information about any of the sex hormones and DNA methylation was available for 302 males and 254 females. Among the females, 28 females were pre-menopausal at time of the sex hormone assessment. We excluded premenopausal women and those who reported use of hormone therapy (N=4) or were missing information on hormone therapy at either visit (N=0). This left 204 postmenopausal women without hormone therapy for the analysis.

Invecchiare in Chianti, aging in the Chianti area (InCHIANTI)

Subjects and study

The InCHIANTI study is population-based study studying aging among individuals from the Chianti region in Italy (Invecchiare in Chianti, aging in the Chianti area).[8] The observations were collected at baseline in 1998. All participants provided written informed consent to participate in this study. The study complied with the Declaration of Helsinki and The Italian National Institute of Research and Care on Aging Institutional Review Board approved the study protocol.

CpG methylation status was estimated using the Illumina Infinium HumanMethylation450 BeadChip (Illumina Inc., San Diego, CA). Methylation beta values were generated using the Bioconductor minfi package with Noob background correction. More details regarding the DNA methylation were previously published.[3]

The details for the sex hormone measurement have also been previously described in details.[9] Briefly, fasting blood samples were drawn between 7 and 8 AM and were stored at −80°C until the analysis. Estradiol was measured by ultrasensitive RIA (DSL-4800 Diagnostic Systems Laboratories, Webster, TX), while total testosterone and DHEAS were assayed using the commercial kits from Diagnostic Systems Laboratories, Webster, TX. Finally, SHBG was measured by a radioimmunoassay from Diagnostic Products Corporation, Los Angeles, CA.[9]

For men, the lowest concentrations measured for these sex hormones were 2.35 pg/ml, 15.23 nmol/L, 20 ng/dl, and 4 ug/dl for estradiol, SHBG, testosterone, and DHEAS respectively. For women, the lowest concentrations measured for these sex hormones were 1 pg/ml, 26.88 nmol/L, 1 ng/dl, and 4.07 ug/dl for estradiol, SHBG, testosterone, and DHEAS, respectively.

Among the InCHIANTI, the population was of European descent. There were 793 individuals of European/White race. Information about any of the sex hormones and DNA methylation was available for 222 males and 258 females. Among the females, 39 females were pre-menopausal at time of the sex hormone assessment. We excluded premenopausal women and those who reported use of hormone therapy (N=1) or were missing information on hormone therapy at either visit (N=3). This left 193 postmenopausal women without hormone therapy for the analysis.
