## Supplemental figures descriptions for "Higher testosterone and testosterone/estradiol ratio in men are associated with better epigenetic estimators of mortality risk"

**Supplemental figure 1.** Scatterplots for the various sex steroid hormones concentrations by age and study, stratified by sex. Figure 1a: Females estrone concentration; 1b: Females estradiol concentration; 1c: Females SHBG concentration; 1d: Females Testosterone concentration; 1e: Females TE ratio; 1f: Females DHEAS concentration; 1g: Males estrone concentration; 1h: Males estradiol concentration; 1i: Males SHBG concentration; 1j: Males Testosterone concentration; 1k: Males TE ratio; 1l: Males DHEAS concentration.

**Supplemental figure 2.**

Correlations between the various sex hormones, including bioavailable sex hormones, stratified by sex (with men in the upper/right quadrant and women in the lower/left quadrant)

**Supplemental figure 3.**

Correlations between the various DNA methylation biomarkers, stratified by sex (with men in the upper/right quadrant and women in the lower/left quadrant)
