## Supplementary figures and images for "Higher testosterone and testosterone/estradiol ratio in men are associated with better epigenetic estimators of mortality risk"

### supplemental figure 1

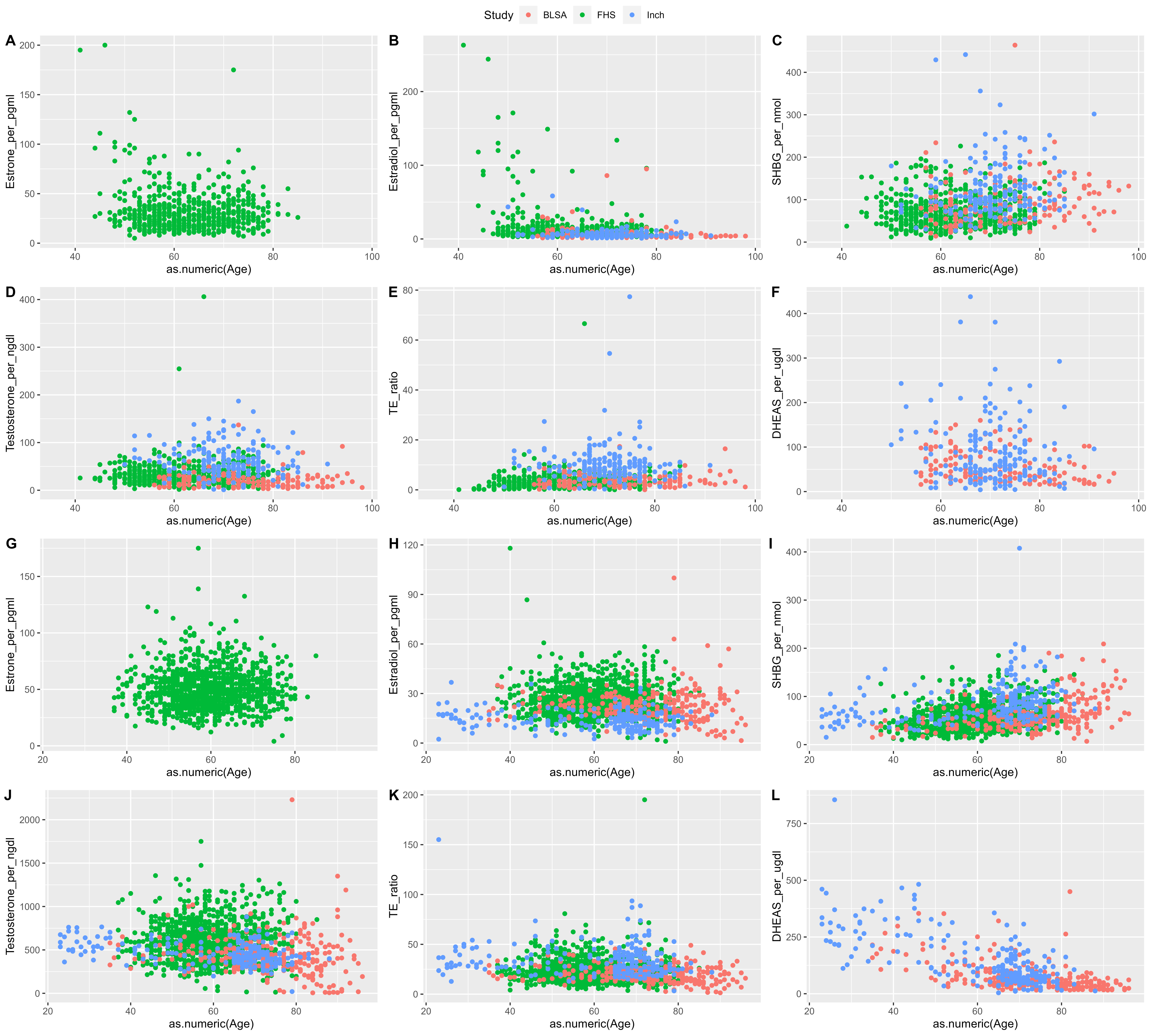

### supplemental figure 2

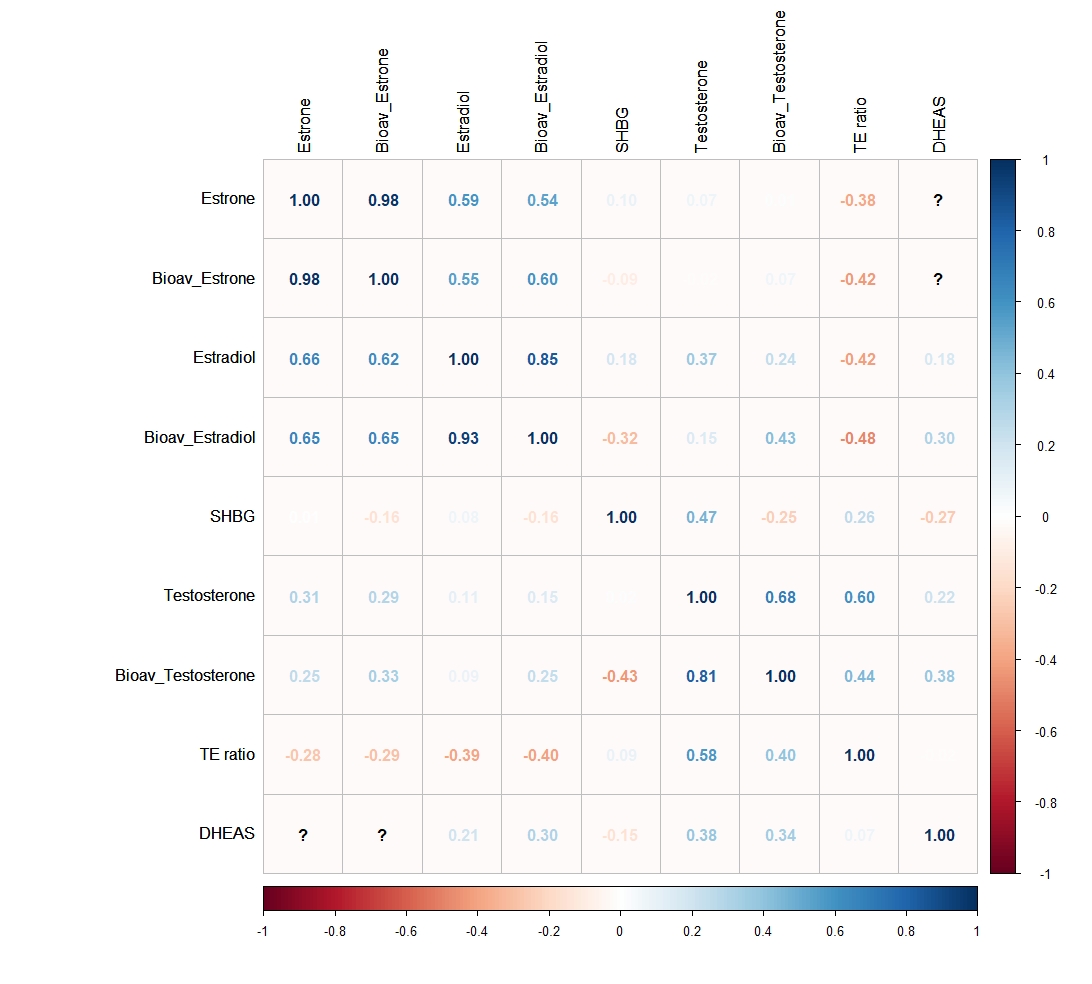

### supplemental figure 3

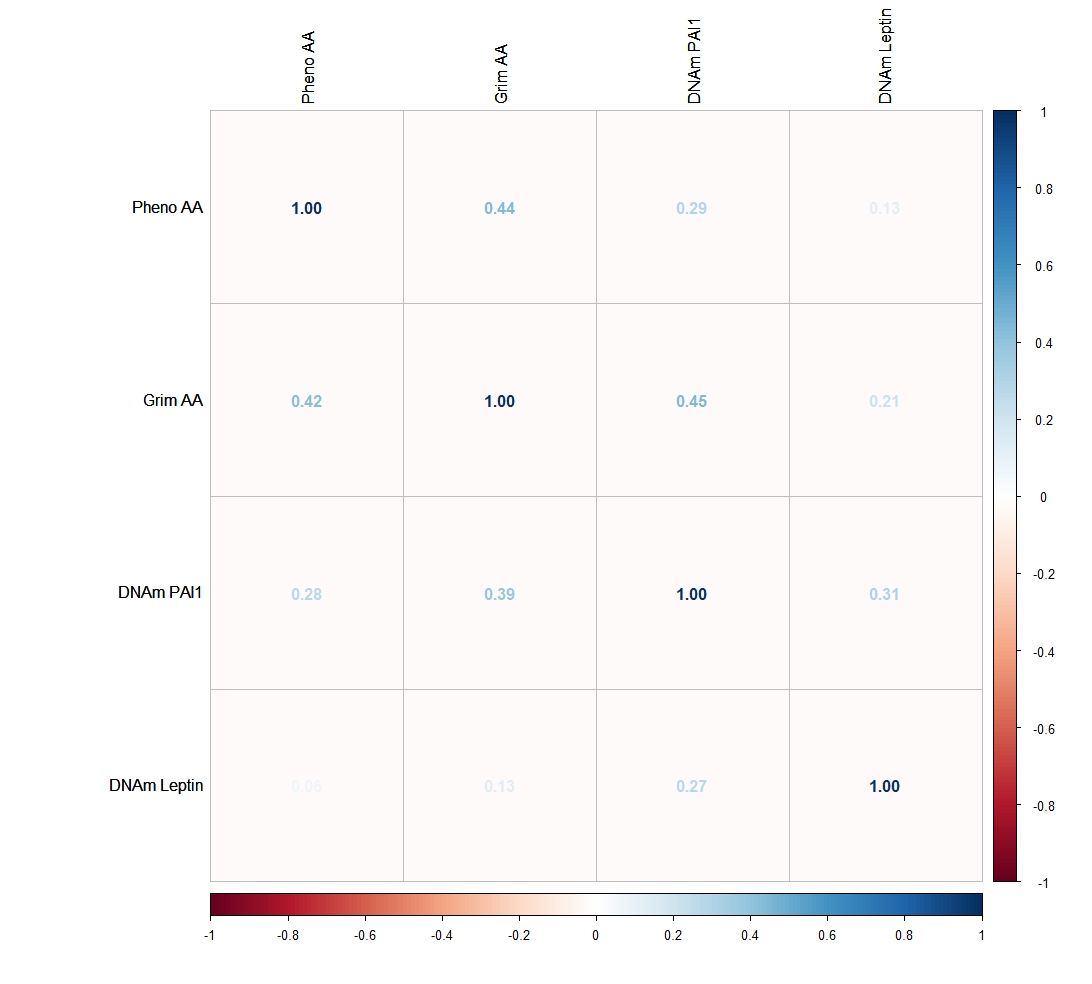
